# Prevalence and associated factors of meeting minimum dietary diversity among pregnant and non-pregnant women of reproductive age in three sub-Saharan African countries

**DOI:** 10.64898/2026.08.24.26361193

**Authors:** Mai-Lei Woo Kinshella, Marie-Laure Volvert, Angela Koech, Hawanatu Jah, Anifa Vala, Marleen Temmerman, Anna Roca, Umberto D’Alessandro, Esperança Sevene, Marianne Vidler, Akshdeep Sandhu, Jeffrey N Bone, Sarka Lisonkova, Laura A. Magee, Peter von Dadelszen, Rajavel Elango, Sophie E. Moore, the PRECISE Network

## Abstract

**Introduction:** Food insecurity and undernutrition persist in much of sub-Saharan Africa. Women of reproductive age (WRA) who fail to meet the minimum dietary diversity (MDD-W) have inadequate nutrient intakes and increased risk of adverse pregnancy outcomes. This study assessed MDD-W in The Gambia, Kenya, and Mozambique and identified determinants.

**Methods:** A food list-based 24-hour recall was conducted within the PRECISE Network, a prospective cohort study with pregnant and non-pregnant WRA in The Gambia, Kenya, and Mozambique. We descriptively summarized dietary diversity scores and rates of MDD-W (≥5 out of 10 food groups) and very low dietary diversity (≤2 food groups). We evaluated associated factors (demographic/household characteristics, socio-economic status, women’s autonomy), using multivariable regression models performed on R Studio (version 4.2.3).

**Results:** Dietary intake data from 7,715 women (1,846 from The Gambia, 3,209 from Kenya, 2,660 from Mozambique) showed that 47.7% met MDD-W (65.1% The Gambia, 45.0% Kenya, 39.2% Mozambique). Pregnant women had a slightly higher rate of meeting MDD-W compared with non-pregnant WRA (48.4% pregnant vs 45.6% non-pregnant [aOR 1.65, 95% CI: 1.40, 1.95]). Higher educational attainment, professional and small business occupations, pregnancy status, parity, household size, marital status and being from The Gambia were protective factors for meeting MDD-W. Poverty and living alone were risk factors for unmet MDD-W. Poverty and country of residence (Mozambique), were risk factors for very low dietary diversity.

**Conclusion:** A majority of the PRECISE cohort did not meet MDD-W, including both pregnant and non-pregnant WRA, suggesting inadequate micronutrient status before pregnancy and limited dietary diversity improvement during pregnancy. Socio-economic indicators are key determinants of adequate dietary diversity, but local contextualisation is essential. Our study highlights the importance of nutrition-specific and-sensitive interventions in women and girls across the lifespan.

## Introduction

An estimated 277 million people in sub-Saharan Africa were undernourished in 2023, equating to 23.2% of the population, with higher rates in East Africa (28.6%) compared with West (16.0%) and Southern Africa (9.6%)[1]. Women are vulnerable to gender inequities in poverty, and become additionally burdened with higher nutritional requirements during pregnancy and lactation [2–5]. Eating a variety of foods is a key component of diet quality and higher dietary diversity is associated with better micronutrient adequacy [6–8]. Almost all micronutrients must be obtained from the diet and are critical to human health and development, particularly during pregnancy, where micronutrient deficiencies have been associated with pregnancy loss, preterm delivery, small birth size, birth defects and metabolic abnormalities [9].

A Sustainable Development Goal 2 indicator for Ending Hunger, the minimum dietary diversity for women (MDD-W) indicator was developed by the Food and Agricultural Organization of the United Nations (FAO) to summarize micronutrient adequacy among women of reproductive age (WRA) [10,11]. MDD-W The MDD-W assessments among pregnant and non-pregnant WRA in sub-Saharan Africa have reported low dietary diversity, with rates of unmet MDD-W typically between 40-80% of the population [12–22], though some report as high as 97.2% [23]. However, understanding MDD-W in sub-Saharan Africa is challenged by limited coverage across the continent, with a majority of studies from Ethiopia [15,16,19,24–29] and relatively fewer studies conducted in Southern [13,14] and West Africa [17,21,22,30]. Qualitative research from The Gambia, Kenya, and Mozambique found that community members identified diverse dietary patterns as nutritious for pregnant women, but described barriers in practice [31]. This study assessed the prevalence of meeting MDD-W among women in the PRECISE (PREgnancy Care Integrating translational Science Everywhere) Network prospective cohort study in The Gambia, Kenya, and Mozambique and factors associated with dietary diversity.

## Methods

### Study design, setting, and participants

This study is part of the PRECISE Network prospective cohort study, with recruitment occurring 19 June 2019 to 22 December 2021 in The Gambia, 2 July 2019 to 16 November 2021 in Mozambique, and 24 June 2019 to 31 December 2022 in Kenya [32]. The PRECISE Network (**Table S1**) comprehensively collected social, demographic, nutritional, clinical and biological data to situate women within their local contexts. The study was conducted on the North Bank of The Gambia, and in Kilifi County in the coastal region of Kenya, and Manhiça District in the southern region of Mozambique, for representation from West, East, and Southern Africa, respectively. PRECISE primarily recruited pregnant women presenting at health facilities, with a smaller sample of non-pregnant WRA presenting at the same health facilities recruited. Detailed description of the PRECISE Network cohort is reported in Craik *et al*., [32].

### Study variables and outcome

The dietary diversity score (DDS) is the number of healthy food groups consumed, ranging from 0 (no food groups consumed) to 10 (all food groups consumed). The ten MDD-W main food groups include: (i) grains, white roots and tubers, and plantains, (ii) pulses, (iii) nuts and seeds, (iv) milk and milk products, (v) meat, poultry and fish, (vi) eggs, (vii) dark green leafy vegetables, (viii) other vitamin A-rich fruits and vegetables, (ix) other vegetables, and (x) other fruits. Intakes of less than 15g (approximately a tablespoon for most foods), were not counted towards a food group, following the FAO guideline [33].

For the current sub-analysis of the PRECISE cohort, meeting the MDD-W was the primary study outcome. MDD-W is a simple, food-based indicator validated for resource-limited settings to assess adequate dietary diversity. Meeting minimum diversity is a dichotomous indicator (yes/no) defined as consuming at least five food groups in the previous 24-hours [33]. A very low dietary diversity threshold of two or less food groups consumed was explored as a secondary outcome.

Participant characteristics were retrieved from the PRECISE Network database. Basic demographic variables included: country of residence (The Gambia, Kenya, Mozambique), cohort (pregnant or non-pregnant WRA), residence (rural, peri-urban, urban), age (15-19 years olds, 20-24 years old, 25-34 years, ≥35 years old), parity (nulliparous or parous), and marital status (married or co-habiting with partner, single or never married, separated or divorced). Socio-economic status (SES) variables included education (highest school level attained: primary school, secondary school, higher education), occupation (housewife, entrepreneurial/informal work such as small business or market trader, labourer in large scale agriculture, factory, and construction, student, professional), poverty (Poverty Probability Index Score, categorized into quintiles ranging from Q1 least likelihood of poverty to Q5 highest likelihood of poverty). Autonomy variables included decision-maker for household spending (husband/partner, father/father-in-law, mother/mother-in-law, participant herself, other) and women’s autonomy for decision-making in household spending (yes/no). Household characteristics included gender of household money decision-maker (male-or female-headed household, household composition (living with husband/partner, living in an extended household with in-law family, living with natal family including parents and/or relatives without husband/partner, and living alone or only with participant’s children) and household size (small household 1-3 people, medium household 4-6 people, large household 7-9 people). Male-headed households were defined as the husband, male partner, father, father-in-law, brother or brother-in-law for the household spending decision-maker. Female-headed households were defined as the participant’s mother, mother-in-law, sister, sister-in-law, or participant herself for the household spending decision-maker. All demographic characteristics were self-reported.

### Data collection and analysis

Trained data collectors administered face-to-face PRECISE Network surveys after obtaining written informed consent, which included collecting demographics, dietary data, and medical histories over four visits during gestation and postpartum. For the assessment of MDD-W, women responded yes or no to consuming local foods corresponding to ten food groups in the previous 24 hours [33].

Descriptive statistics summarized demographic characteristics, DDS, and the proportions that met MDD-W by country and overall. As an ordinal food group diversity score [11,34], median DDS along with interquartile ranges (IQR) were reported. Multivariable logistic regression models were performed to evaluate protective and risk factors associated with meeting MDD-W and risk factors for very low dietary diversity. These were reported as odds ratios (ORs) with their corresponding 95% confidence intervals. The model was adjusted for basic demographics (country of residence, pregnant/non-pregnant status, rural/periurban/urban residence, age at enrollment, parity, marital status), socio-economic status (highest education attained, occupation, poverty index quintile), women’s autonomy (decision-making for household spending), and household composition (gender of primary decision-maker for household spending, living arrangements and household size). These were informed by a literature review of dietary diversity studies from sub-Saharan Africa, including age [12,13,15,23], rural residency [14,17], parity [19], woman’s educational status [12,15–17,19,23,35–37], women’s occupation/employment status [37], women’s autonomy in decision-making [15,35], gender of household head [12,13,19,25,30] and household size [12,13,19,25,30]. Additionally, factors were informed by qualitative research within study communities [31]. Multicollinearity was assessed using variance inflation factor (VIF), with VIF values above 5 removed from the model [38]. No variables were removed due to multicollinearity. All analyses were performed on R Studio (version 4.2.3).

### Ethical considerations

The study received ethics approval from the University of British Columbia (H20-00143). Approval for the PRECISE study was also obtained in King’s College London (Ref HR-17/18-7855), Aga Khan University Hospital (Ref 2018/REC-74), The Gambia Government/The Medical Research Council, The Gambia Joint committee (Ref SCC 1619) and the Mozambique Ministry of Health, National Bioethics Committee for Health (545/CNBS/18). Participants gave informed consent to participate in the study before taking part. Participant confidentiality was protected using codes to de-identify data.

### Patient and public involvement

In addition to community engagement activities in the wider PRECISE Network [32], qualitative research was conducted with women, family members and community opinion leaders regarding factors that influenced maternal dietary diversity [31].

## Results

### Participant characteristics

Of the 8,757 women from PRECISE, complete dietary intake data were collected for a total of N = 7,715 women (The Gambia N=1,846; Kenya N= 3,209; Mozambique N=2,660) (**Fig 1**). There were 1,549 non-pregnant women of reproductive age and 6,166 pregnant women included in the analyses.

**Fig 1.**
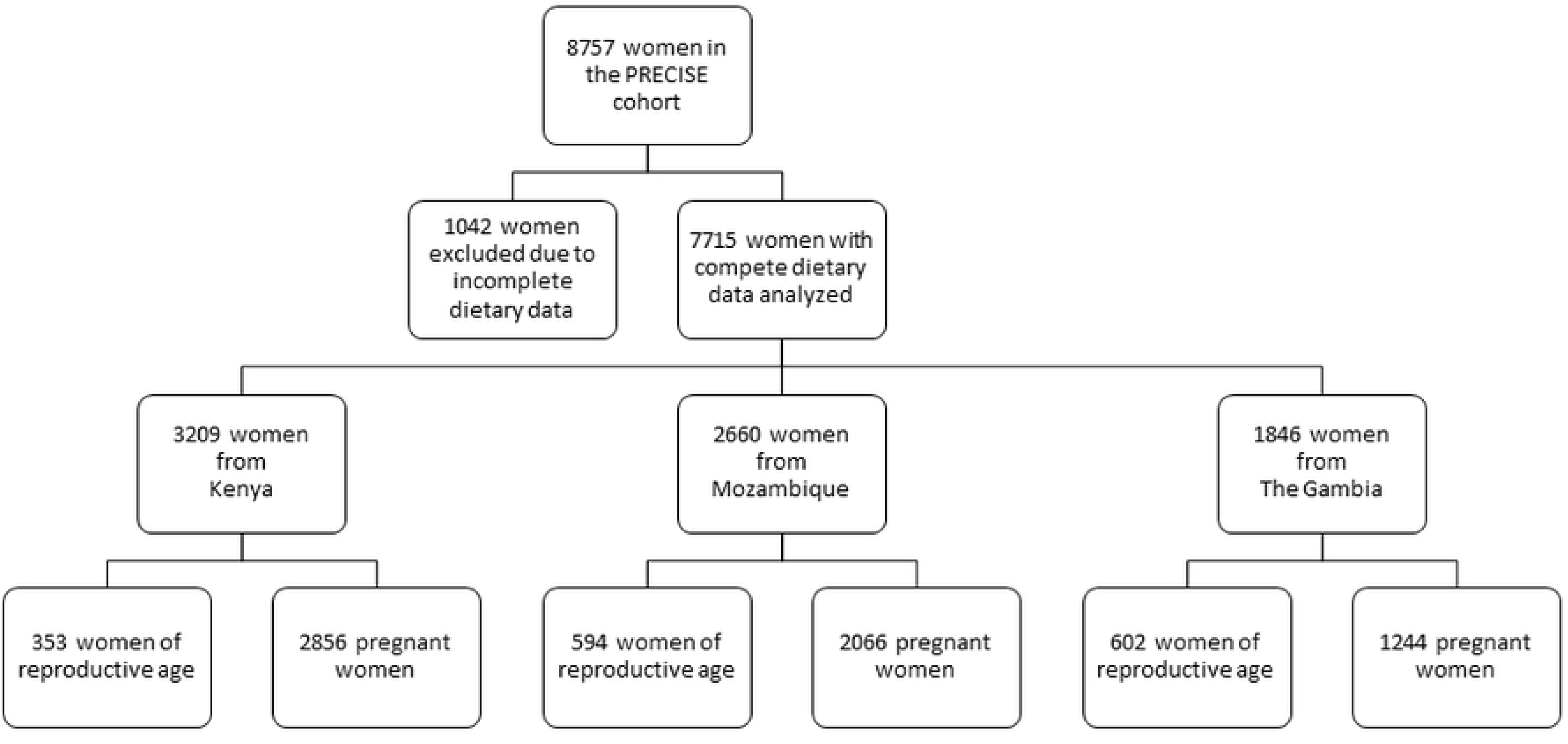
Profile of PRECISE participants

Overall, a little over half (53%) the women lived in urban areas. However, in The Gambia, a higher proportion lived in rural areas (63%) (**Table 1**). Most women across the three countries were parous (72.4%) and married or co-habiting with their partner (78.6%). There was an overall median age of 26 years [IQR 21, 31]. Primary school education was reported by 37.6% of women in the cohort, while 34.3% reported secondary school and 6.8% reported higher education. Many women from The Gambia did not report attending any formal schooling (63.0 %). Two-thirds of women reported being a housewife as their occupation (66.6%). There was an overall 21.4% likelihood of being below the poverty line in the cohort, with higher likelihood of poverty among women from The Gambia. Most households were male-headed (79.0%), husband/partners were most frequently the decision-maker for household spending (66.6%) and most women did not have autonomy to make decisions about household spending on her own (87.8%). Women most frequently reported living with their husband or partner (46.7%), and, in The Gambia, this was often within an extended household with in-laws (55.6%). There was a median of 5 people [IQR 3,9] in a household, with Kenyan participants more frequently reporting smaller household sizes and Gambian participants more frequently reporting larger household sizes.

**Table 1.**
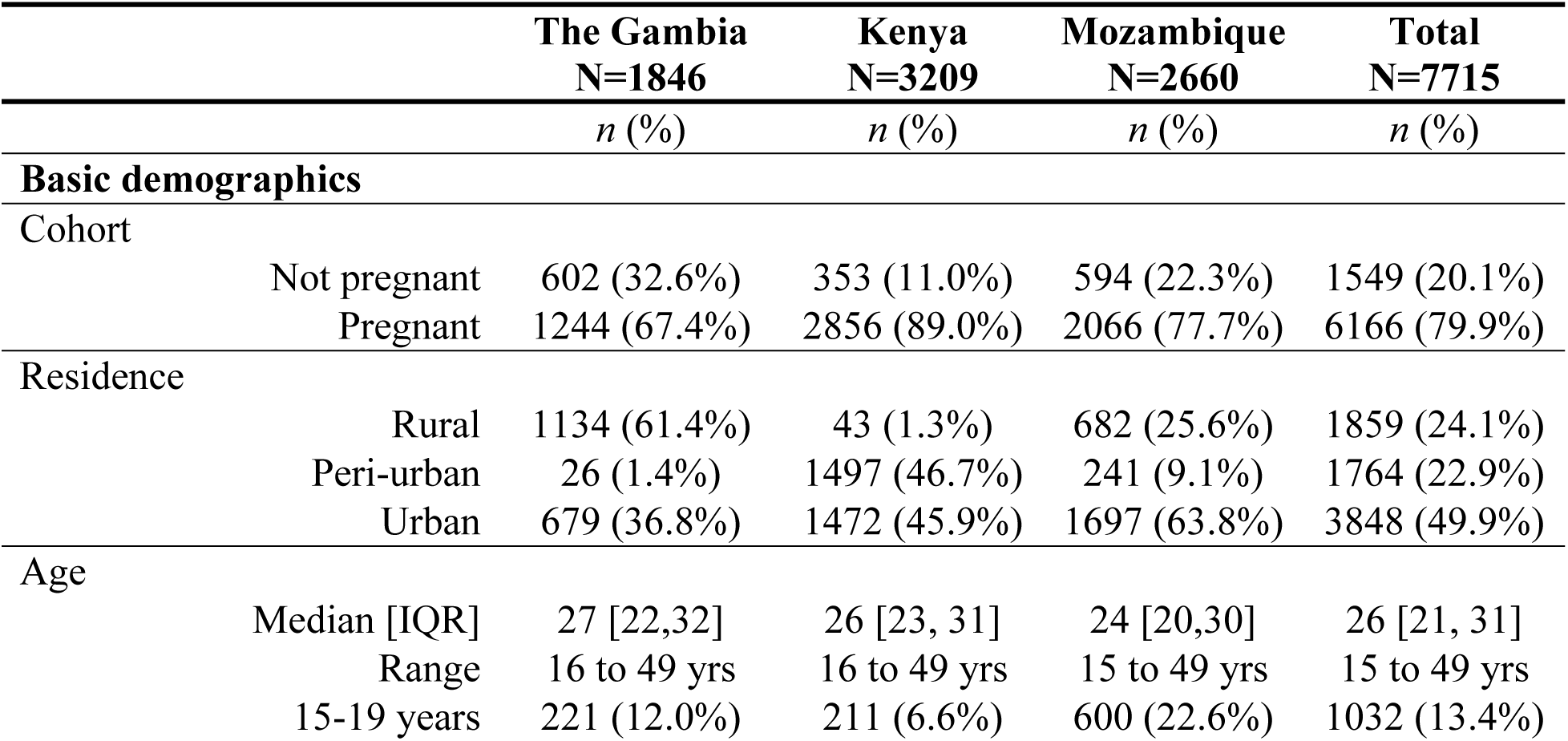
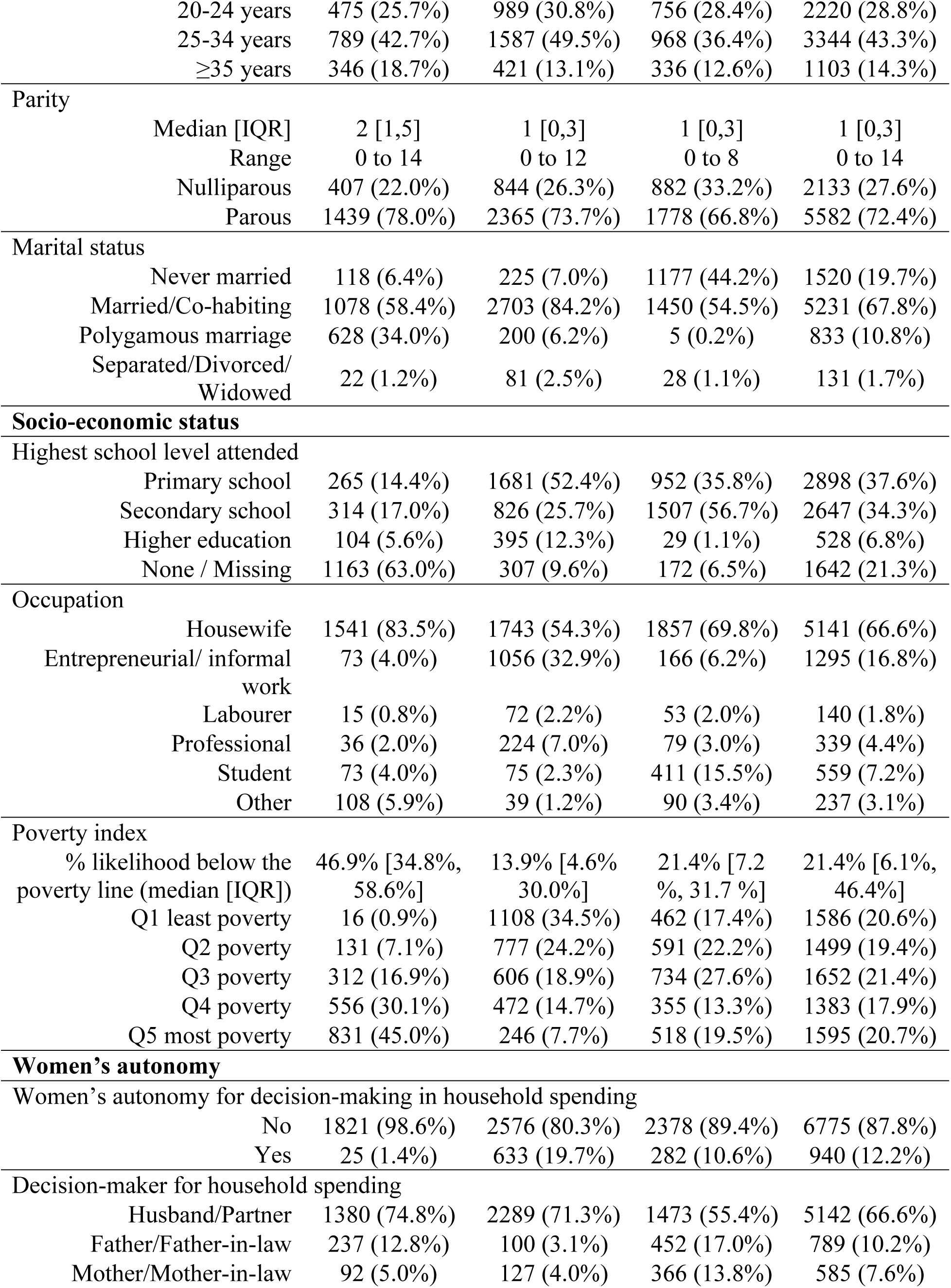
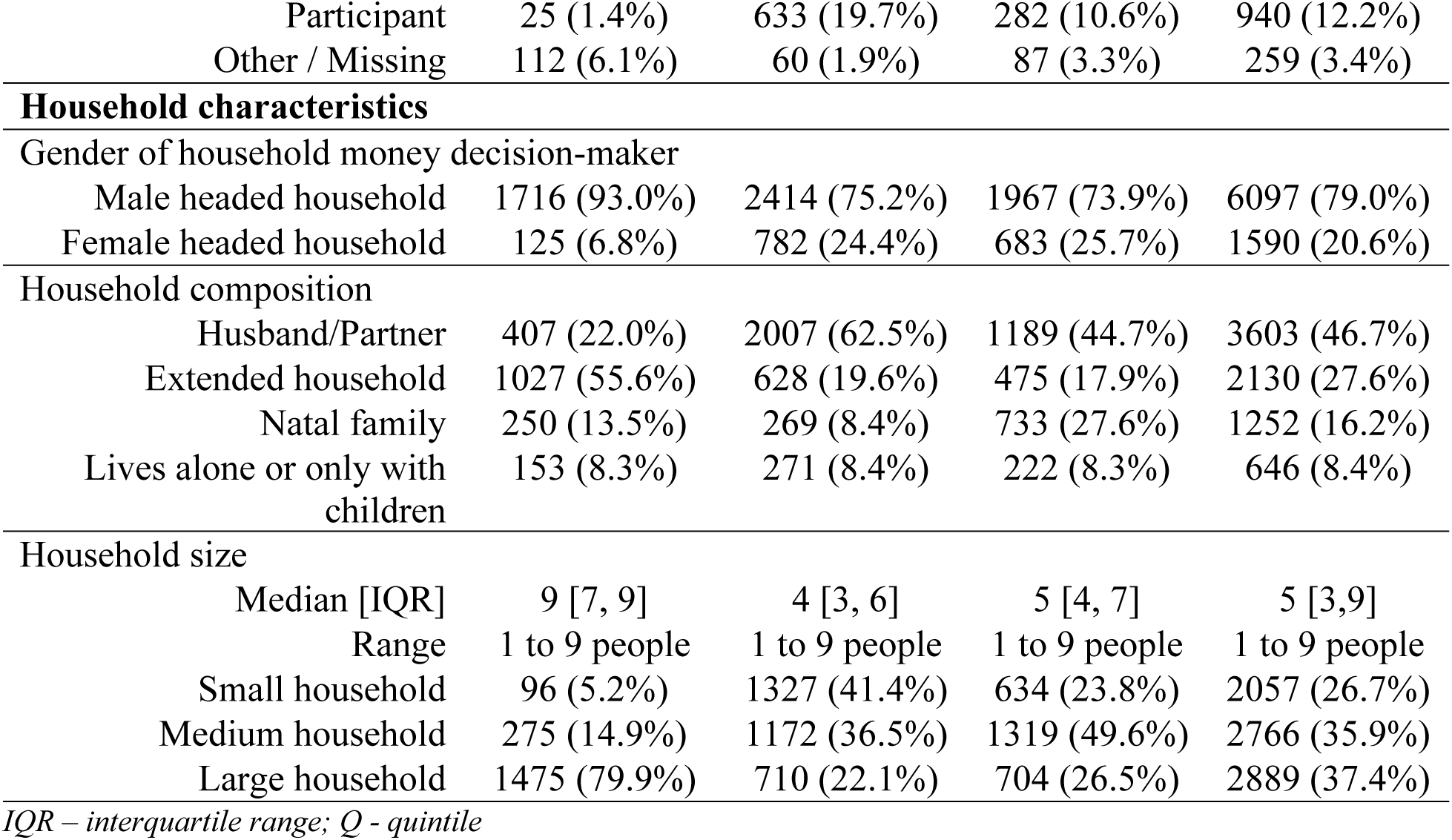
Participant characteristics.

|  |  | **The Gambia**<br>**N=1846** | **Kenya**<br>**N=3209** | **Mozambique**<br>**N=2660** | **Total**<br>**N=7715** |
| --- | --- | --- | --- | --- | --- |
|  |  | *n (%)* | *n (%)* | *n (%)* | *n (%)* |
| **Basic demographics** |  |  |  |  |  |
| Cohort |  |  |  |  |  |
|  | Not pregnant | 602 (32.6%) | 353 (11.0%) | 594 (22.3%) | 1549 (20.1%) |
|  | Pregnant | 1244 (67.4%) | 2856 (89.0%) | 2066 (77.7%) | 6166 (79.9%) |
| Residence |  |  |  |  |  |
|  | Rural | 1134 (61.4%) | 43 (1.3%) | 682 (25.6%) | 1859 (24.1%) |
|  | Peri-urban | 26 (1.4%) | 1497 (46.7%) | 241 (9.1%) | 1764 (22.9%) |
|  | Urban | 679 (36.8%) | 1472 (45.9%) | 1697 (63.8%) | 3848 (49.9%) |
| Age |  |  |  |  |  |
|  | Median [IQR] | 27 [22,32] | 26 [23, 31] | 24 [20,30] | 26 [21, 31] |
|  | Range | 16 to 49 yrs | 16 to 49 yrs | 15 to 49 yrs | 15 to 49 yrs |
|  | 15-19 years | 221 (12.0%) | 211 (6.6%) | 600 (22.6%) | 1032 (13.4%) |
|  | 20-24 years | 475 (25.7%) | 989 (30.8%) | 756 (28.4%) | 2220 (28.8%) |
|  | 25-34 years | 789 (42.7%) | 1587 (49.5%) | 968 (36.4%) | 3344 (43.3%) |
|  | ≥35 years | 346 (18.7%) | 421 (13.1%) | 336 (12.6%) | 1103 (14.3%) |
| **Parity** |  |  |  |  |  |
|  | Median [IQR] | 2 [1,5] | 1 [0,3] | 1 [0,3] | 1 [0,3] |
|  | Range | 0 to 14 | 0 to 12 | 0 to 8 | 0 to 14 |
|  | Nulliparous | 407 (22.0%) | 844 (26.3%) | 882 (33.2%) | 2133 (27.6%) |
|  | Parous | 1439 (78.0%) | 2365 (73.7%) | 1778 (66.8%) | 5582 (72.4%) |
| **Marital status** |  |  |  |  |  |
|  | Never married | 118 (6.4%) | 225 (7.0%) | 1177 (44.2%) | 1520 (19.7%) |
|  | Married/Co-habiting | 1078 (58.4%) | 2703 (84.2%) | 1450 (54.5%) | 5231 (67.8%) |
|  | Polygamous marriage | 628 (34.0%) | 200 (6.2%) | 5 (0.2%) | 833 (10.8%) |
|  | Separated/Divorced/<br>Widowed | 22 (1.2%) | 81 (2.5%) | 28 (1.1%) | 131 (1.7%) |
| **Socio-economic status** |  |  |  |  |  |
| **Highest school level attended** |  |  |  |  |  |
|  | Primary school | 265 (14.4%) | 1681 (52.4%) | 952 (35.8%) | 2898 (37.6%) |
|  | Secondary school | 314 (17.0%) | 826 (25.7%) | 1507 (56.7%) | 2647 (34.3%) |
|  | Higher education | 104 (5.6%) | 395 (12.3%) | 29 (1.1%) | 528 (6.8%) |
|  | None / Missing | 1163 (63.0%) | 307 (9.6%) | 172 (6.5%) | 1642 (21.3%) |
| **Occupation** |  |  |  |  |  |
|  | Housewife | 1541 (83.5%) | 1743 (54.3%) | 1857 (69.8%) | 5141 (66.6%) |
|  | Entrepreneurial/ informal<br>work | 73 (4.0%) | 1056 (32.9%) | 166 (6.2%) | 1295 (16.8%) |
|  | Labourer | 15 (0.8%) | 72 (2.2%) | 53 (2.0%) | 140 (1.8%) |
|  | Professional | 36 (2.0%) | 224 (7.0%) | 79 (3.0%) | 339 (4.4%) |
|  | Student | 73 (4.0%) | 75 (2.3%) | 411 (15.5%) | 559 (7.2%) |
|  | Other | 108 (5.9%) | 39 (1.2%) | 90 (3.4%) | 237 (3.1%) |
| **Poverty index** |  |  |  |  |  |
|  | % likelihood below the<br>poverty line (median [IQR]) | 46.9% [34.8%,<br>58.6%] | 13.9% [4.6%<br>30.0%] | 21.4% [7.2<br>%, 31.7 %] | 21.4% [6.1%,<br>46.4%] |
|  | Q1 least poverty | 16 (0.9%) | 1108 (34.5%) | 462 (17.4%) | 1586 (20.6%) |
|  | Q2 poverty | 131 (7.1%) | 777 (24.2%) | 591 (22.2%) | 1499 (19.4%) |
|  | Q3 poverty | 312 (16.9%) | 606 (18.9%) | 734 (27.6%) | 1652 (21.4%) |
|  | Q4 poverty | 556 (30.1%) | 472 (14.7%) | 355 (13.3%) | 1383 (17.9%) |
|  | Q5 most poverty | 831 (45.0%) | 246 (7.7%) | 518 (19.5%) | 1595 (20.7%) |
| **Women's autonomy** |  |  |  |  |  |
| **Women's autonomy for decision-making in household spending** |  |  |  |  |  |
|  | No | 1821 (98.6%) | 2576 (80.3%) | 2378 (89.4%) | 6775 (87.8%) |
|  | Yes | 25 (1.4%) | 633 (19.7%) | 282 (10.6%) | 940 (12.2%) |
| **Decision-maker for household spending** |  |  |  |  |  |
|  | Husband/Partner | 1380 (74.8%) | 2289 (71.3%) | 1473 (55.4%) | 5142 (66.6%) |
|  | Father/Father-in-law | 237 (12.8%) | 100 (3.1%) | 452 (17.0%) | 789 (10.2%) |
|  | Mother/Mother-in-law | 92 (5.0%) | 127 (4.0%) | 366 (13.8%) | 585 (7.6%) |

|  |  |  |  |  |
| --- | --- | --- | --- | --- |
| Participant | 25 (1.4%) | 633 (19.7%) | 282 (10.6%) | 940 (12.2%) |
| Other / Missing | 112 (6.1%) | 60 (1.9%) | 87 (3.3%) | 259 (3.4%) |
| **Household characteristics** |  |  |  |  |
| Gender of household money decision-maker |  |  |  |  |
| Male headed household | 1716 (93.0%) | 2414 (75.2%) | 1967 (73.9%) | 6097 (79.0%) |
| Female headed household | 125 (6.8%) | 782 (24.4%) | 683 (25.7%) | 1590 (20.6%) |
| Household composition |  |  |  |  |
| Husband/Partner | 407 (22.0%) | 2007 (62.5%) | 1189 (44.7%) | 3603 (46.7%) |
| Extended household | 1027 (55.6%) | 628 (19.6%) | 475 (17.9%) | 2130 (27.6%) |
| Natal family | 250 (13.5%) | 269 (8.4%) | 733 (27.6%) | 1252 (16.2%) |
| Lives alone or only with children | 153 (8.3%) | 271 (8.4%) | 222 (8.3%) | 646 (8.4%) |
| Household size |  |  |  |  |
| Median [IQR] | 9 [7, 9] | 4 [3, 6] | 5 [4, 7] | 5 [3,9] |
| Range | 1 to 9 people | 1 to 9 people | 1 to 9 people | 1 to 9 people |
| Small household | 96 (5.2%) | 1327 (41.4%) | 634 (23.8%) | 2057 (26.7%) |
| Medium household | 275 (14.9%) | 1172 (36.5%) | 1319 (49.6%) | 2766 (35.9%) |
| Large household | 1475 (79.9%) | 710 (22.1%) | 704 (26.5%) | 2889 (37.4%) |

*IQR – interquartile range; Q - quintile*

### Dietary diversity

Women in the PRECISE cohort had a median DDS of four [IQR 3, 6], with the full range from zero to ten food groups consumed in the previous 24 hours reported (**Fig S1, Fig S2**). Median DDS of four was found across participant and household characteristics, except Gambian participants, rural women, those with higher education, labourer or professional occupation, least poverty (Q1), and those living in extended and large households, which had a median DDS of five (**Table 2**).

**Table 2.**
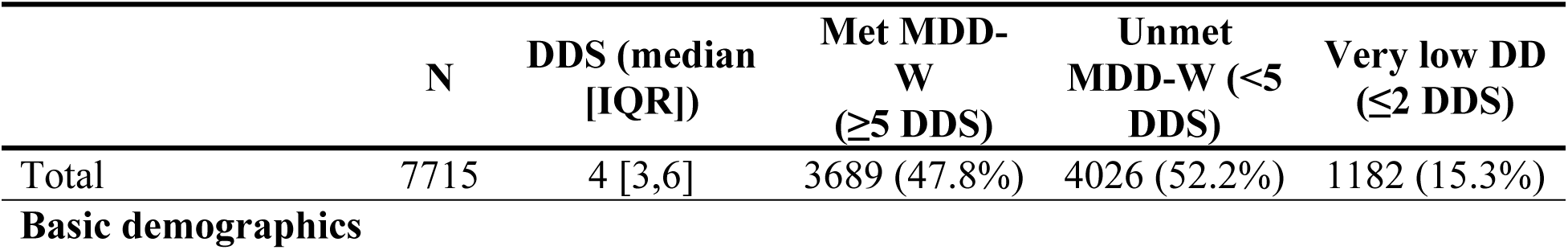
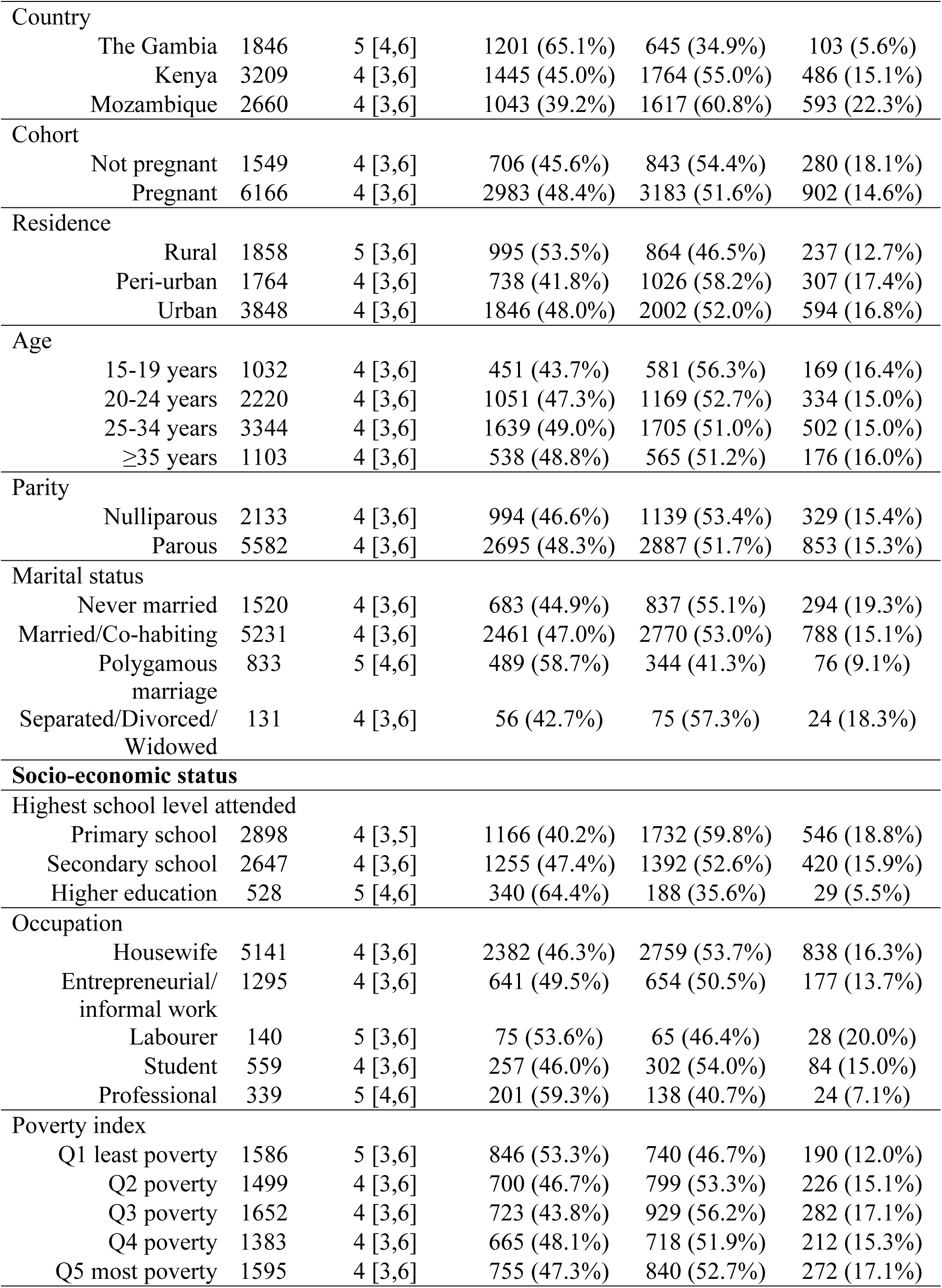
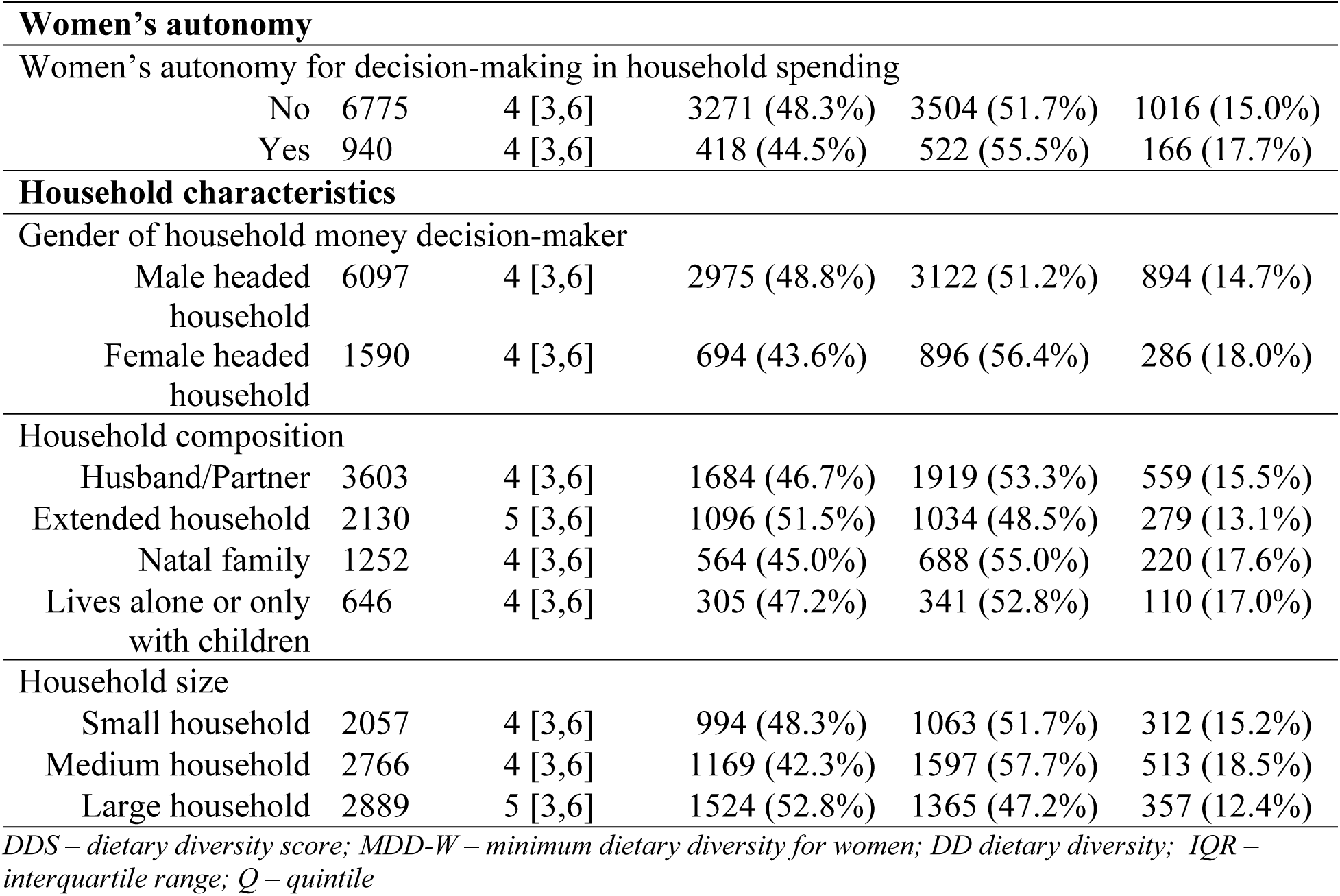
Dietary diversity scores and prevalences.

|  | N | DDS (median [IQR]) | Met MDD-W (≥5 DDS) | Unmet MDD-W (<5 DDS) | Very low DD (≤2 DDS) |
| --- | --- | --- | --- | --- | --- |
| Total | 7715 | 4 [3,6] | 3689 (47.8%) | 4026 (52.2%) | 1182 (15.3%) |
| **Basic demographics** |  |  |  |  |  |
| Country |  |  |  |  |  |
| The Gambia | 1846 | 5 [4,6] | 1201 (65.1%) | 645 (34.9%) | 103 (5.6%) |
| Kenya | 3209 | 4 [3,6] | 1445 (45.0%) | 1764 (55.0%) | 486 (15.1%) |
| Mozambique | 2660 | 4 [3,6] | 1043 (39.2%) | 1617 (60.8%) | 593 (22.3%) |
| Cohort |  |  |  |  |  |
| Not pregnant | 1549 | 4 [3,6] | 706 (45.6%) | 843 (54.4%) | 280 (18.1%) |
| Pregnant | 6166 | 4 [3,6] | 2983 (48.4%) | 3183 (51.6%) | 902 (14.6%) |
| Residence |  |  |  |  |  |
| Rural | 1858 | 5 [3,6] | 995 (53.5%) | 864 (46.5%) | 237 (12.7%) |
| Peri-urban | 1764 | 4 [3,6] | 738 (41.8%) | 1026 (58.2%) | 307 (17.4%) |
| Urban | 3848 | 4 [3,6] | 1846 (48.0%) | 2002 (52.0%) | 594 (16.8%) |
| Age |  |  |  |  |  |
| 15-19 years | 1032 | 4 [3,6] | 451 (43.7%) | 581 (56.3%) | 169 (16.4%) |
| 20-24 years | 2220 | 4 [3,6] | 1051 (47.3%) | 1169 (52.7%) | 334 (15.0%) |
| 25-34 years | 3344 | 4 [3,6] | 1639 (49.0%) | 1705 (51.0%) | 502 (15.0%) |
| ≥35 years | 1103 | 4 [3,6] | 538 (48.8%) | 565 (51.2%) | 176 (16.0%) |
| Parity |  |  |  |  |  |
| Nulliparous | 2133 | 4 [3,6] | 994 (46.6%) | 1139 (53.4%) | 329 (15.4%) |
| Parous | 5582 | 4 [3,6] | 2695 (48.3%) | 2887 (51.7%) | 853 (15.3%) |
| Marital status |  |  |  |  |  |
| Never married | 1520 | 4 [3,6] | 683 (44.9%) | 837 (55.1%) | 294 (19.3%) |
| Married/Co-habiting | 5231 | 4 [3,6] | 2461 (47.0%) | 2770 (53.0%) | 788 (15.1%) |
| Polygamous marriage | 833 | 5 [4,6] | 489 (58.7%) | 344 (41.3%) | 76 (9.1%) |
| Separated/Divorced/<br>Widowed | 131 | 4 [3,6] | 56 (42.7%) | 75 (57.3%) | 24 (18.3%) |
| **Socio-economic status** |  |  |  |  |  |
| Highest school level attended |  |  |  |  |  |
| Primary school | 2898 | 4 [3,5] | 1166 (40.2%) | 1732 (59.8%) | 546 (18.8%) |
| Secondary school | 2647 | 4 [3,6] | 1255 (47.4%) | 1392 (52.6%) | 420 (15.9%) |
| Higher education | 528 | 5 [4,6] | 340 (64.4%) | 188 (35.6%) | 29 (5.5%) |
| Occupation |  |  |  |  |  |
| Housewife | 5141 | 4 [3,6] | 2382 (46.3%) | 2759 (53.7%) | 838 (16.3%) |
| Entrepreneurial/<br>informal work | 1295 | 4 [3,6] | 641 (49.5%) | 654 (50.5%) | 177 (13.7%) |
| Labourer | 140 | 5 [3,6] | 75 (53.6%) | 65 (46.4%) | 28 (20.0%) |
| Student | 559 | 4 [3,6] | 257 (46.0%) | 302 (54.0%) | 84 (15.0%) |
| Professional | 339 | 5 [4,6] | 201 (59.3%) | 138 (40.7%) | 24 (7.1%) |
| Poverty index |  |  |  |  |  |
| Q1 least poverty | 1586 | 5 [3,6] | 846 (53.3%) | 740 (46.7%) | 190 (12.0%) |
| Q2 poverty | 1499 | 4 [3,6] | 700 (46.7%) | 799 (53.3%) | 226 (15.1%) |
| Q3 poverty | 1652 | 4 [3,6] | 723 (43.8%) | 929 (56.2%) | 282 (17.1%) |
| Q4 poverty | 1383 | 4 [3,6] | 665 (48.1%) | 718 (51.9%) | 212 (15.3%) |
| Q5 most poverty | 1595 | 4 [3,6] | 755 (47.3%) | 840 (52.7%) | 272 (17.1%) |
| **Women's autonomy** |  |  |  |  |  |
| Women's autonomy for decision-making in household spending |  |  |  |  |  |
| No | 6775 | 4 [3,6] | 3271 (48.3%) | 3504 (51.7%) | 1016 (15.0%) |
| Yes | 940 | 4 [3,6] | 418 (44.5%) | 522 (55.5%) | 166 (17.7%) |
| **Household characteristics** |  |  |  |  |  |
| Gender of household money decision-maker |  |  |  |  |  |
| Male headed household | 6097 | 4 [3,6] | 2975 (48.8%) | 3122 (51.2%) | 894 (14.7%) |
| Female headed household | 1590 | 4 [3,6] | 694 (43.6%) | 896 (56.4%) | 286 (18.0%) |
| Household composition |  |  |  |  |  |
| Husband/Partner | 3603 | 4 [3,6] | 1684 (46.7%) | 1919 (53.3%) | 559 (15.5%) |
| Extended household | 2130 | 5 [3,6] | 1096 (51.5%) | 1034 (48.5%) | 279 (13.1%) |
| Natal family | 1252 | 4 [3,6] | 564 (45.0%) | 688 (55.0%) | 220 (17.6%) |
| Lives alone or only with children | 646 | 4 [3,6] | 305 (47.2%) | 341 (52.8%) | 110 (17.0%) |
| Household size |  |  |  |  |  |
| Small household | 2057 | 4 [3,6] | 994 (48.3%) | 1063 (51.7%) | 312 (15.2%) |
| Medium household | 2766 | 4 [3,6] | 1169 (42.3%) | 1597 (57.7%) | 513 (18.5%) |
| Large household | 2889 | 5 [3,6] | 1524 (52.8%) | 1365 (47.2%) | 357 (12.4%) |

DDs – dietary diversity score; MDD-W – minimum dietary diversity for women; DD dietary diversity; IQR – interquartile range; Q – quintile

Overall, 47.8% across the cohort met MDD-W, including 65.1% of women from The Gambia, 45.0% of women from Kenya, and 39.2% of women from Mozambique (**Table 2**). Although pregnant and non-pregnant women both had a median DDS of four [IQR 3, 6], pregnant women had a slightly higher proportion of meeting MDD-W (48.4% pregnant vs 45.6% non-pregnant). Very low dietary diversity was reported by 15.3% of the cohort.

### Factors associated with meeting minimum dietary diversity for women

Country of residence was associated with meeting MDD-W, with three-fold higher odds of adequate dietary diversity among Gambian women (aOR 3.28, 95% CI: 2.57, 4.19), compared with Kenyan women (**Table 3**). Compared with non-pregnant WRA, pregnant women had higher odds of meeting MDD-W (aOR 1.65, 95% CI: 1.40, 1.95). Across the cohort, parous women had higher odds of meeting MDD-W (aOR 1.18, 95% CI: 1.02, 1.38), compared with nulliparous women in the adjusted analyses. Marital status was also associated with meeting MDD-W, with higher odds of adequate dietary diversity among never married women (aOR 1.49, 95% CI: 1.21, 1.83) compared with those who were married and cohabiting. Mozambique country of residence, peri-urban/urban residency, adolescent age or older adulthood, polygamous marriage, and separated/divorced/widowed marital status had wide confidence intervals in the adjusted analyses, indicating uncertain associations with MDD-W status.

**Table 3.**
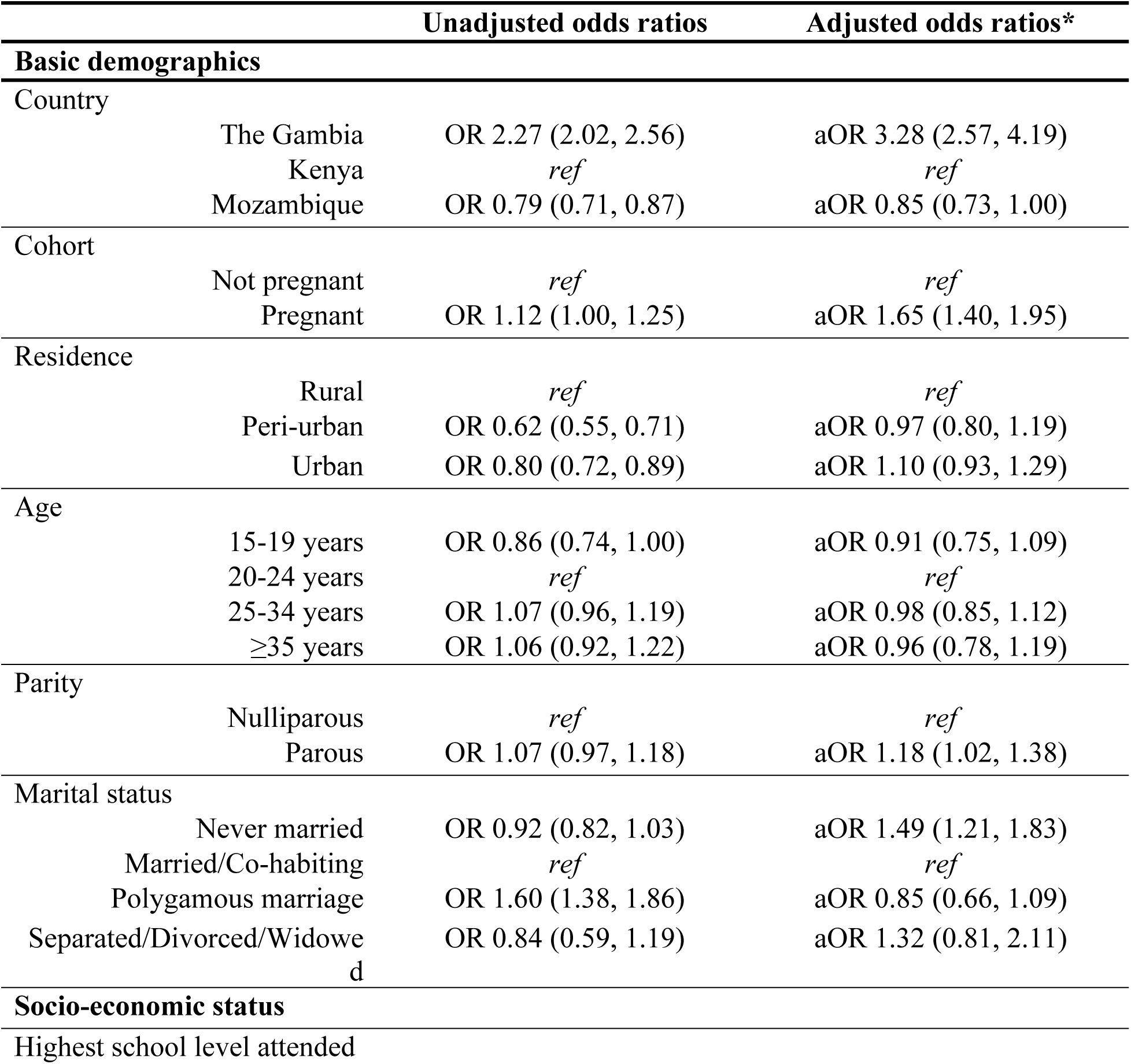
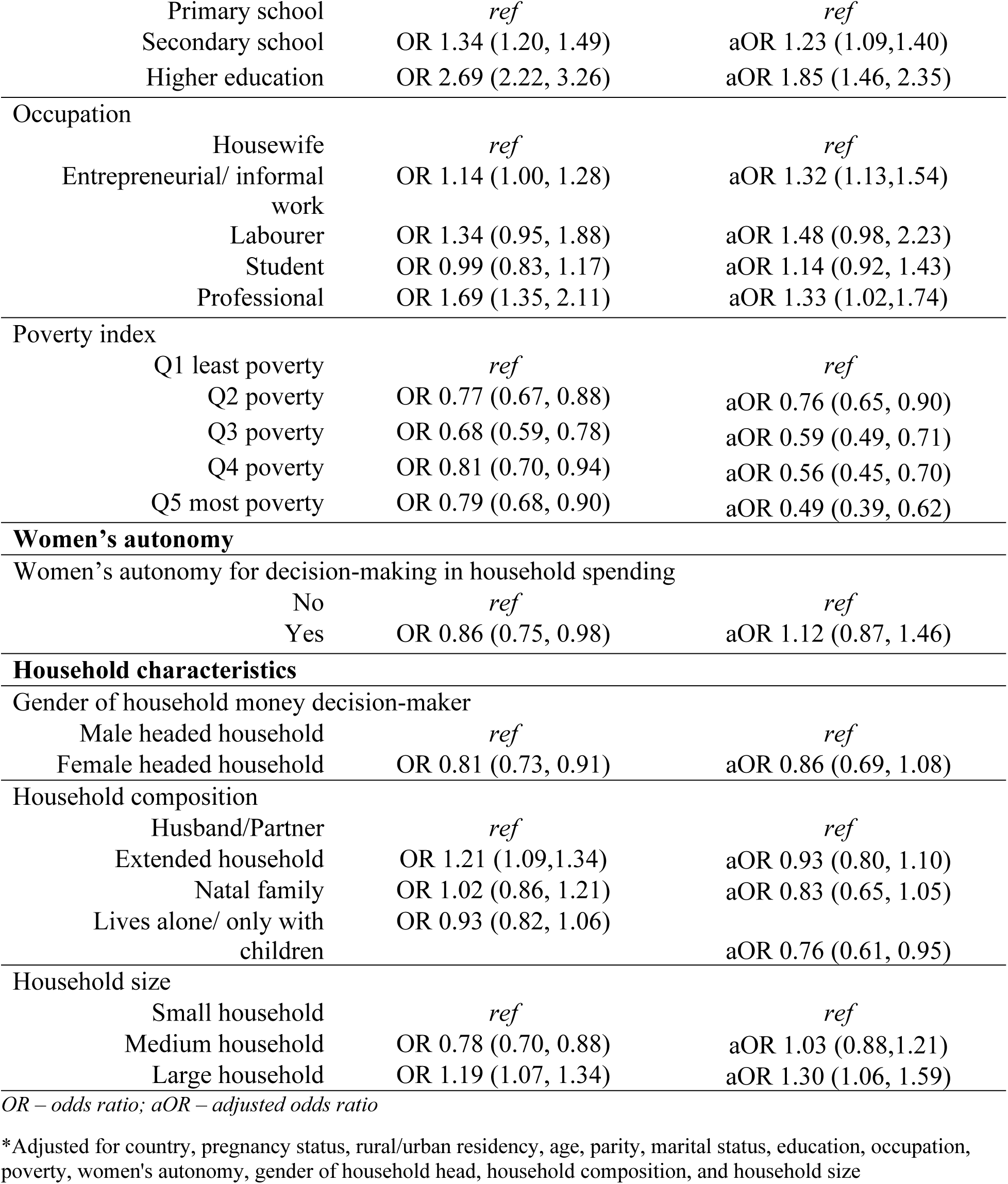
Factors associated with meeting MDD-W.

|  |  | Unadjusted odds ratios | Adjusted odds ratios* |
| --- | --- | --- | --- |
| **Basic demographics** |  |  |  |
| Country |  |  |  |
|  | The Gambia | OR 2.27 (2.02, 2.56) | aOR 3.28 (2.57, 4.19) |
|  | Kenya | *ref* | *ref* |
|  | Mozambique | OR 0.79 (0.71, 0.87) | aOR 0.85 (0.73, 1.00) |
| Cohort |  |  |  |
|  | Not pregnant | *ref* | *ref* |
|  | Pregnant | OR 1.12 (1.00, 1.25) | aOR 1.65 (1.40, 1.95) |
| Residence |  |  |  |
|  | Rural | *ref* | *ref* |
|  | Peri-urban | OR 0.62 (0.55, 0.71) | aOR 0.97 (0.80, 1.19) |
|  | Urban | OR 0.80 (0.72, 0.89) | aOR 1.10 (0.93, 1.29) |
| Age |  |  |  |
|  | 15-19 years | OR 0.86 (0.74, 1.00) | aOR 0.91 (0.75, 1.09) |
|  | 20-24 years | *ref* | *ref* |
|  | 25-34 years | OR 1.07 (0.96, 1.19) | aOR 0.98 (0.85, 1.12) |
|  | ≥35 years | OR 1.06 (0.92, 1.22) | aOR 0.96 (0.78, 1.19) |
| Parity |  |  |  |
|  | Nulliparous | *ref* | *ref* |
|  | Parous | OR 1.07 (0.97, 1.18) | aOR 1.18 (1.02, 1.38) |
| Marital status |  |  |  |
|  | Never married | OR 0.92 (0.82, 1.03) | aOR 1.49 (1.21, 1.83) |
|  | Married/Co-habiting | *ref* | *ref* |
|  | Polygamous marriage | OR 1.60 (1.38, 1.86) | aOR 0.85 (0.66, 1.09) |
|  | Separated/Divorced/Widowed | OR 0.84 (0.59, 1.19) | aOR 1.32 (0.81, 2.11) |
| **Socio-economic status** |  |  |  |
| Highest school level attended |  |  |  |

|  |  |  |
| --- | --- | --- |
| Primary school | *ref* | *ref* |
| Secondary school | OR 1.34 (1.20, 1.49) | aOR 1.23 (1.09,1.40) |
| Higher education | OR 2.69 (2.22, 3.26) | aOR 1.85 (1.46, 2.35) |
| **Occupation** |  |  |
| Housewife | *ref* | *ref* |
| Entrepreneurial/ informal work | OR 1.14 (1.00, 1.28) | aOR 1.32 (1.13,1.54) |
| Labourer | OR 1.34 (0.95, 1.88) | aOR 1.48 (0.98, 2.23) |
| Student | OR 0.99 (0.83, 1.17) | aOR 1.14 (0.92, 1.43) |
| Professional | OR 1.69 (1.35, 2.11) | aOR 1.33 (1.02,1.74) |
| **Poverty index** |  |  |
| Q1 least poverty | *ref* | *ref* |
| Q2 poverty | OR 0.77 (0.67, 0.88) | aOR 0.76 (0.65, 0.90) |
| Q3 poverty | OR 0.68 (0.59, 0.78) | aOR 0.59 (0.49, 0.71) |
| Q4 poverty | OR 0.81 (0.70, 0.94) | aOR 0.56 (0.45, 0.70) |
| Q5 most poverty | OR 0.79 (0.68, 0.90) | aOR 0.49 (0.39, 0.62) |
| **Women's autonomy** |  |  |
| Women's autonomy for decision-making in household spending |  |  |
| No | *ref* | *ref* |
| Yes | OR 0.86 (0.75, 0.98) | aOR 1.12 (0.87, 1.46) |
| **Household characteristics** |  |  |
| Gender of household money decision-maker |  |  |
| Male headed household | *ref* | *ref* |
| Female headed household | OR 0.81 (0.73, 0.91) | aOR 0.86 (0.69, 1.08) |
| Household composition |  |  |
| Husband/Partner | *ref* | *ref* |
| Extended household | OR 1.21 (1.09,1.34) | aOR 0.93 (0.80, 1.10) |
| Natal family | OR 1.02 (0.86, 1.21) | aOR 0.83 (0.65, 1.05) |
| Lives alone/ only with children | OR 0.93 (0.82, 1.06) | aOR 0.76 (0.61, 0.95) |
| Household size |  |  |
| Small household | *ref* | *ref* |
| Medium household | OR 0.78 (0.70, 0.88) | aOR 1.03 (0.88,1.21) |
| Large household | OR 1.19 (1.07, 1.34) | aOR 1.30 (1.06, 1.59) |

*OR – odds ratio; aOR – adjusted odds ratio*
\*Adjusted for country, pregnancy status, rural/urban residency, age, parity, marital status, education, occupation, poverty, women's autonomy, gender of household head, household composition, and household size

All three SES indicators were associated with MDD-W status. Compared with women who self-reported being a housewife, entrepreneurial small business and informal work (aOR 1.32, 95% CI: 1.13,1.54) and professional occupations (aOR 1.33, 95% CI: 1.02,1.74) had higher odds of meeting MDD-W. Poverty was inversely associated with meeting MDD-W, with lower odds of adequate dietary diversity with increasing likelihood of poverty. A dose response was also evident with education; in comparison with women who attended primary school, secondary school attendance (aOR 1.23, 95% CI: 1.09,1.40) and higher education (aOR 1.85, 95% CI: 1.46, 2.35) had higher odds of meeting MDD-W.

Household characteristics associated with MDD-W status included lower odds of meeting MDD-W among women who reported living alone or only with their children (aOR 0.76, 95% CI: 0.61, 0.95); women who lived in extended households with their in-laws or those who lived with their parents or relatives did not substantially differ in their rates of meeting MDD-W compared with those who lived with their husbands and partners. We also found higher odds of meeting MDD-W among larger households (aOR 1.30, 95% CI: 1.06, 1.59), compared with small household with three people or less. Neither women’s autonomy for household spending nor gender of household head appeared associated with MDD-W status.

### Risk factors of very low dietary diversity

Women from Mozambique (aOR 1.54, 95% CI: 1.25, 1.90) and higher poverty were associated with eating two or less food groups in the previous 24 hours (**Table S2**). In particular, the two highest poverty quintiles had over two-fold higher odds of very low dietary diversity compared with the lowest poverty quintile (Q5 aOR 2.41, 95%CI: 1.78, 3.28); Q4 aOR 2.08, 95%CI: 1.55, 2.79). Factors associated with dietary diversity are summarized in **Fig 2**.

**Fig 2.**
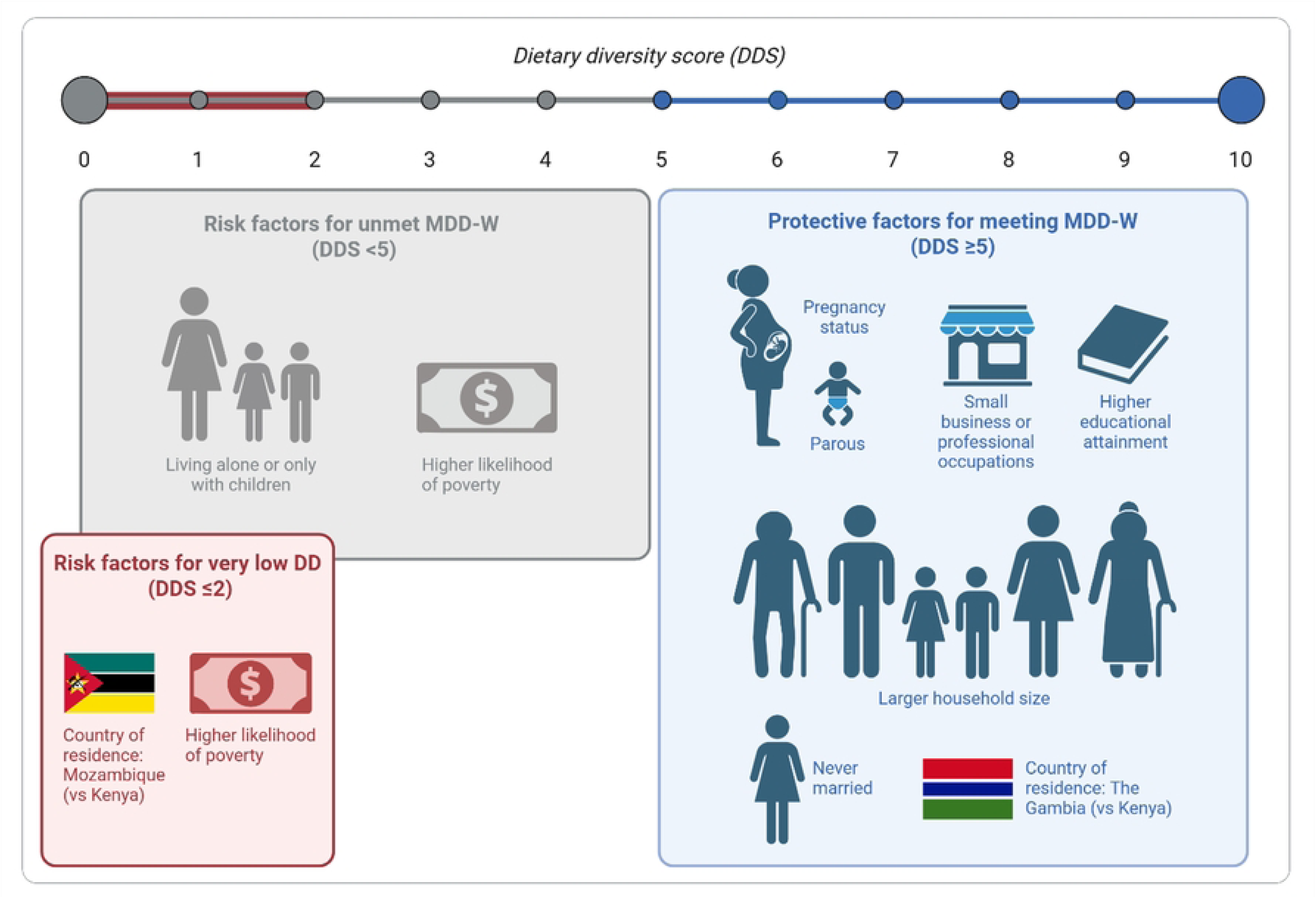
Risk and protective factors for dietary diversity in the PRECISE Network

## Discussion

### Summary of findings and comparisons to the literature

Most women in the PRECISE cohort did not achieve adequate dietary diversity, as assessed by the MDD-W, and almost one in six women reported low diversity, consuming two or less food groups in the previous 24-hours. Country of residence and socio-economic indicators emerged as key factors of dietary diversity, both for MDD-W and as factors associated with heightened vulnerability to very low dietary diversity. Being from Mozambique emerged as a risk factor particularly for low dietary diversity. Being from The Gambia, pregnancy status, parity, never married marital status, higher educational attainment, small business and professional occupations, and larger household size emerged as potentially protective factors to support higher dietary diversity.

Our overall prevalence of meeting MDD-W at 47.7% of women in the PRECISE cohort is in line with a systematic review from Ethiopia that found a pooled rate of 41% [16]. Our country-specific rates were lower than previously reported for Kenya (45.0% in PRECISE vs 69%) and Mozambique (39.2% in PRECISE vs 52%) [39], and no other published MDD-W data from The Gambia were found. This may be influenced by seasonal and regional differences within each country as well as longer periods of data collection in PRECISE (The Gambia Jun 2019-Dec 2022 PRECISE vs Kenya Jul-Aug 2021/ Mozambique Nov 2021 – Jan 2022 [39]). A recent study from Uganda found that rates of unmet MDD-W appear to be increasing [40] and the 2024 The State of Food Security and Nutrition in the World report described stabilizing rates of undernutrition globally except in the African region where rates have continued to climb [1]. This underscores the importance of understanding underlying factors.

SES variables, including women’s education, occupation, and likelihood of poverty, emerged as strong influencing factors of dietary diversity. Larger effect sizes for higher education compared to secondary education and increasing likelihood of poverty are suggestive of dose responses. Higher educational attainment and household wealth were the most frequently reported indicators of meeting MDD-W among studies conducted in sub-Saharan Africa [12–17,19,21–23,25,30,35–37,40], though definitions varied between studies. Perhaps due to various occupational categories being compared, some previous studies found a significant association between women’s employment status and higher dietary diversity [37,41] and others not [17,20,35,42–45]. In addition to professional occupations where women may receive a salary, our findings also highlight the potential for small business and market trading work to support dietary diversity. This is inline with a qualitative study from Nigeria that found economic empowerment supportive of dietary diversity, particularly to purchase non-staple food items [46]. Variations in measurement and local contexts may contribute to the heterogeneity in findings across the literature.

### Dietary diversity in context

Our study adds to the literature by highlighting regional variability in dietary diversity status across West, East, and Southern Africa, and evaluating dietary diversity among pregnant and non-pregnant WRA within in the same communities, timeframe and dietary assessment methodology. The comprehensive PRECISE Network database complemented by qualitative maternal diet research helped to provide nuance on SES indicators. For example, rates of meeting MDD-W were higher among Gambian women even though higher rates of no formal education, higher poverty levels, and lower rates of paid occupations were reported from The Gambia compared with Kenya and Mozambique. Higher dietary diversity may be influenced by local diets and meals as Gambian food often consisted of many ingredients cooked together in a pot, compared with typical meals in Kenya and Mozambique [31]. Overall, pregnancy status was associated with likelihood of meeting MDD-W, even after controlling for SES and other variables. This reinforced qualitative findings that diverse diets were valued for pregnant women in PRECISE study communities [31]. However, persistent challenges in practice shared by community members [31] is reflected in continued high rate of inadequate dietary diversity (51.6%) among pregnant women.

Perhaps due to systematic gender inequities that may disadvantage female-headed households within patriarchal societies [47], we did not find associations for women’s autonomy or gender of household primary decision-maker on meeting MDD-W. However, we did find higher odds of adequate dietary diversity among never married women compared with those who were married and cohabiting, which appears to be largely driven by women from Mozambique, where many men in the region migrate to South Africa for work, and single women from rural Southern Mozambique have high rates of engaging in paid employment [48]. Previous studies of indicators of women’s empowerment have been mixed with reports of significant [15,35,41] and non-significant associations with dietary diversity [43,49,50]. Gender of household head in previous studies have either favoured male-headed households [12,18] or reported non-significant associations with women’s dietary diversity [13,35,37,41]. Overall, our study found that women living alone or only with their children had a lower likelihood of meeting MDD-W, which has not been previously evaluated as a determinant of MDD-W to the best of our knowledge.

### Implications for research and policy

High rates of unmet dietary diversity in the PRECISE cohort suggests substantial vulnerabilities to inadequate micronutrient status, which threatens the wellbeing of both mother and offspring. Our findings, in alignment with other reports of low MDD-W rates in Africa, is a call to action to strengthen nutrition in women and girls across the lifespan [51,52]. Nutrition-specific interventions such as supplementation, fortification and dietary counselling can be strengthened during antenatal care and expanded alongside regular screening for anaemia and undernutrition within family planning and reproductive health initiatives. Investing in dietary diverse and nutritious school feeding programs can support the dietary status of adolescent girls, along with boys. Nutrition-sensitive interventions, including promoting education and women’s economic and social empowerment, may also improve MDD-W rates, as out study found strong associations between MDD-W status and SES indicators. Currently, sub-optimal prevalence of meeting MDD-W among non-pregnant women suggests that many women are not starting their pregnancies with adequate micronutrient status. While being pregnant was associated with higher odds of adequate dietary diversity, continued sub-optimal prevalence of achieving MDD among pregnant women highlights that dietary diversity did not substantially improve during pregnancy.

However, there was a range in dietary diversity scores highlighting variation within the population. Our findings support the need for both nutrition-specific interventions, such as raising awareness of maternal dietary diversity among families and health professionals, alongside nutrition-sensitive interventions, such as strengthening education and employment initiatives for women and girls and reducing poverty [53–56]. Additionally, with the understanding that low dietary diversity and subsequent inadequate micronutrient status may heighten risk of adverse pregnancy outcomes, women with unmet MDD-W or very low dietary diversity could also be identified for additional maternity follow-up and support, such as enhanced community health worker and/or community group-based health education and microfinance programs [57–60]. Future research could explore the potential of MDD-W assessments within prenatal care settings as an opportunity for nutritional counselling and as a screening tool to identify vulnerable mothers.

### Strengths and limitations

The current analysis utilized the standardized MDD-W instrument, which has been validated in resource-limited settings, and included non-pregnant WRA and pregnant women from the same communities and time-period in the study cohort to enable comparisons between the two groups. Dietary assessments were conducted across the three countries with women over the span of two and a half years with a large sample size, though there was a pause in recruitment in April-July 2020 due to the COVID-19 pandemic. Strengths include the use of culturally adapted food lists and reduced potential recall bias in the 24-hour recall method compared to other food frequency questionnaires that request recall over longer periods of time. Due to self-reported dietary data, there is potential for measurement error from social desirability bias where participants report consuming food items, they understand to be healthy. Over-reporting of beans and peas, dark green leafy vegetables, vitamin A-rich fruit and vegetables, and other fruits has been previously documented with the MDD-W tool and may overestimate rates of meeting minimum dietary diversity [34]. Furthermore, dietary data was collected over multiple seasons across three and a half years and the impact of seasonality has not been accounted for, which represents a limitation of the study and a recommended area for future research. Potential impact of the COVID-19 pandemic on dietary diversity is also an area that can be further explored.

## Conclusion

Our assessment found 52.3% of women in the PRECISE cohort across The Gambia, Kenya, and Mozambique did not meet MDD-W, suggesting sub-optimal micronutrient adequacy among these populations. Our findings highlight important differences between the African regions studied, such as West Africa compared with East and Southern Africa; and, within populations including by pregnant status, marital status, household composition, and SES, that warrant further investigation. While socio-economic factors emerged as overall key determinants of dietary diversity both in our study and across the literature, factors need to be contextualized to local settings, norms and practices.

## Data Availability

All relevant data are within the paper and its Supporting Information files.

## Acknowledgements

The authors would like to express their gratitude to the PRECISE Team for their support and all the women and their families who participated in the study. This manuscript is part of the PRECISE (PREgnancy Care Integrating translational Science, Everywhere) Network.

## Supporting information

**Table S1.** The PRECISE Network

**Fig S1.** Dietary diversity scores in the PRECISE Network

**Fig S2.** Dietary diversity scores by PRECISE Network country

**Table S2.** Risk factors for very low dietary diversity

